# Vaccine introductions in the WHO African Region, 2023–26: a country-level ecological analysis by Gavi eligibility and conflict-affected status

**DOI:** 10.64898/2026.06.19.26356078

**Authors:** Adidja Amani, Pamela Mitula, Aschalew Teka Bekele, Celestin Danwang, Hermann Didi Ngossaki, Yolande Vuo Masembe, Amavi Edinam Agbenu, Lanzy Achille, Claude Mangobo, Ado Mpia Bwaka, Marcellin Nimpa Mengouo, Mutale Mumba, Terna Nomhwange, Nosheen Safdar, André Arsène Bita Fouda, Alam Khattak, Lynda Rey, Okot Charles Lukoya, Kwasi Nyarko, Bridget Lindsay Farham, Anuj Kapilashrami, Akpaka A. Kalu, Benido Impouma

## Abstract

**Background:** The Immunization Agenda 2030 (IA2030) tracks new and underused vaccine introduction as an access metric, and its mid-term review calls for stronger country ownership, prioritisation, data use and tailored support in conflict-affected and resource-constrained settings; however, national launch status does not measure recurrent financing, implementation, safety or equity. We examined how recent vaccine-programme-change activity was distributed across the WHO African Region.

**Methods:** We conducted a descriptive country-level ecological analysis of all 47 Member States from January 2023 to June 2026. The country was the unit of analysis and contributed one cumulative, unweighted count of nationally endorsed vaccine-programme-change events. Counts were linked to Gavi eligibility, World Bank FY26 conflict-affected status (with a broader fragile and conflict-affected situation [FCS] definition examined in sensitivity analysis) and concurrent system-performance indicators, and modelled with Poisson regression using HC1 robust standard errors. Two Expanded Programme on Immunization (EPI) manager survey waves were summarised at country level. Reporting followed STROBE and RECORD.

**Results:** Seventy-two events were recorded across 38 of 47 Member States: 48 new-antigen introductions, 20 dose or schedule expansions and four combination-vaccine introductions; malaria vaccines accounted for 21. Gavi-eligible conflict-affected countries averaged 2.50 events per country versus 1.27 in both comparison groups. Gavi-eligible conflict-affected status was associated with a higher count (incidence rate ratio [IRR] 1.97, 95% confidence interval [CI] 1.38–2.81; p<0.001), whereas non-Gavi status was not (IRR 1.00, 95% CI 0.54–1.87). The association was strongest for new-antigen introductions (IRR 2.36, 95% CI 1.43–3.91), attenuated but persisted after excluding malaria-vaccine events (IRR 1.77, 95% CI 1.06–2.97), and was robust to baseline introduction-opportunity adjustment (IRR 2.00). Surveys reported constraints in prioritisation, financing, 2026 preparation and post-introduction evaluation.

**Conclusions:** Recent vaccine-programme-change activity concentrated where Gavi eligibility, conflict-affected status and weaker concurrent system performance intersected. The association was attenuated but remained directionally consistent after excluding malaria-vaccine events, indicating that the observed Gavi–conflict pattern was not explained solely by malaria-vaccine rollout. Because the analysis was ecological and country-level, it does not establish causality. Monitoring should pair launch status with financing, implementation, safety, dose-completion, stock-continuity and equity indicators to distinguish policy adoption from sustained protection

**Highlights:**

- 72 programme-change events occurred in 38 of 47 Member States.
- Activity concentrated in Gavi-eligible conflict-affected countries.
- New antigen introductions showed the strongest gradient.
- The gradient attenuated after excluding malaria-vaccine events.
- Launch metrics should be paired with implementation assurance.

## 1. Introduction

Immunisation has been one of the most consequential public-health interventions in the WHO African Region. Over the past five decades, vaccination is estimated to have saved 51.2 million lives in the Region, while global programmes have averted at least 154 million deaths [1,2]. These gains remain incomplete and were placed under pressure by the COVID-19 pandemic: in recent regional reporting, coverage for the third dose of diphtheria–tetanus–pertussis-containing vaccine (DTP3) was 76%, second-dose measles-containing vaccine coverage was 55%, and human papillomavirus (HPV) vaccine coverage among girls was 44%, each below the 90% IA2030 benchmark, while approximately 6·7 million children remained zero-dose [3,4]. New vaccine introduction is therefore occurring not in a stable expansion phase, but amid routine-immunisation recovery, outbreak response and equity pressure.

New and underused vaccine introduction is central to IA2030, which includes a target of at least 500 cumulative introductions in low-income and middle-income countries between 2021 and 2030, and whose mid-term review calls for stronger country ownership, structured prioritisation, subnational data use and tailored support to fragile, conflict-affected and vulnerable settings [1,5]. This target is necessary for monitoring access, but launch status is an incomplete measure of programme performance: a recorded introduction shows that a vaccine has entered a programme; it does not show whether recurrent financing is secured, supply is continuous, health workers are trained, safety surveillance is functional, doses are completed, post-introduction evaluation is conducted, or high-risk populations are equitably reached [6,7]. Political-economy analyses of global health initiatives similarly caution that programme outputs can obscure the country-level practices, resource competition and implementation arrangements generated by external financing architectures [8]. Existing global monitoring describes whether recommended vaccines have entered national schedules; it does not show whether recent programme-change activity is accumulating in countries where financing eligibility, conflict-affected status and weaker system performance intersect.

Previous regional analyses establish that the WHO African Region made substantial progress in vaccine adoption before the present study period. Sambala and colleagues reported that the average number of new and underused vaccines introduced per country increased from three of 12 in 2010 to seven of 12 in 2017, with universal adoption of hepatitis B and Haemophilus influenzae type b vaccines and major expansion of pneumococcal conjugate, rotavirus, inactivated poliovirus and second-dose measles-containing vaccines, and they warned that sustaining these gains would require predictable, reliable financing beyond vaccine purchase [9]. Iwu-Jaja and colleagues extended this baseline to 2022, reporting universal introduction of hepatitis B, Haemophilus influenzae type b and inactivated poliovirus vaccines across all 47 Member States while gaps persisted for hepatitis B birth dose, HPV vaccine and the second dose of inactivated poliovirus vaccine (IPV2) [10]. These studies show that the Region can adopt vaccines at scale; they leave unresolved whether the post-2022 programme-change agenda is distributed according to financing and conflict context, and whether launch-status monitoring captures the recurrent functions required for sustained implementation.

Recent programme changes are operationally heterogeneous and should be appraised against criteria that include disease burden, vaccine effectiveness and safety, opportunity cost, affordability, sustainability and programme capacity [11]. In the regional portfolio these criteria apply differently across products: malaria vaccines require subnational targeting, immunisation–malaria coordination, readiness monitoring and multi-dose completion [12,13]; HPV vaccines require adolescent platforms, school and out-of-school reach, demand generation and sustainable financing [14,15]; hepatitis B birth dose depends on timely maternity and newborn services, where delayed vaccination reduces protection even when eventual coverage improves [16,17]; and schedule expansions, product switches and combination vaccines require procurement, cold-chain, training, safety and recording-system adaptation [18]. A binary introduction indicator therefore collapses operationally different programme changes into a single access measure.

Conflict-affected settings and financing transition add further implementation risk. The World Bank classification distinguishes conflict-affected situations from settings characterised by institutional and social fragility, and is updated annually for strategic and operational decision-making [19,20]. In the analytic classification used here, ten Member States were both Gavi-eligible and listed under the World Bank FY26 conflict-affected category, creating a financing–conflict group in which access opportunity and implementation risk could plausibly coincide. Existing evidence has examined vaccine adoption decisions, single-antigen roll-outs, technical-assistance models and implementation barriers [21–25], but regional evidence remains limited on whether recent activity represents cumulative access progress alone or also a distributional and implementation-assurance challenge.

This study examined vaccine-programme-change activity in the WHO African Region from January 2023 to June 2026. The primary aim was to assess whether recent activity was broadly distributed or patterned by financing and conflict context. Secondary aims were to characterise the portfolio by timing, vaccine or platform and operational type; describe subregional variation; quantify differences in country-level event counts across financing–conflict groups; examine co-occurrence with concurrent system-performance indicators; and identify EPI manager-reported constraints in prioritisation, financing, product-switching and post-introduction evaluation not visible in launch-status monitoring alone.

## 2. Methods

### 2.1 Study design, setting and reporting

We conducted a descriptive country-level ecological analysis of vaccine-programme-change events across all 47 WHO African Region Member States from January 2023 to June 2026. The country was the unit of analysis; each Member State contributed one cumulative event count over the observation window. Countries were grouped using the WHO African Region programme subregional structure: West Africa, Central Africa, and Eastern and Southern Africa. The full country list, subregional grouping, Gavi eligibility and conflict classification are provided in Supplementary Table S3. The study workflow, from data sources to triangulation, is summarised in Figure 1. Reporting followed the STROBE statement and the RECORD extension for routinely collected health data (Supplementary File S1) [26,27].

**Figure 1.**
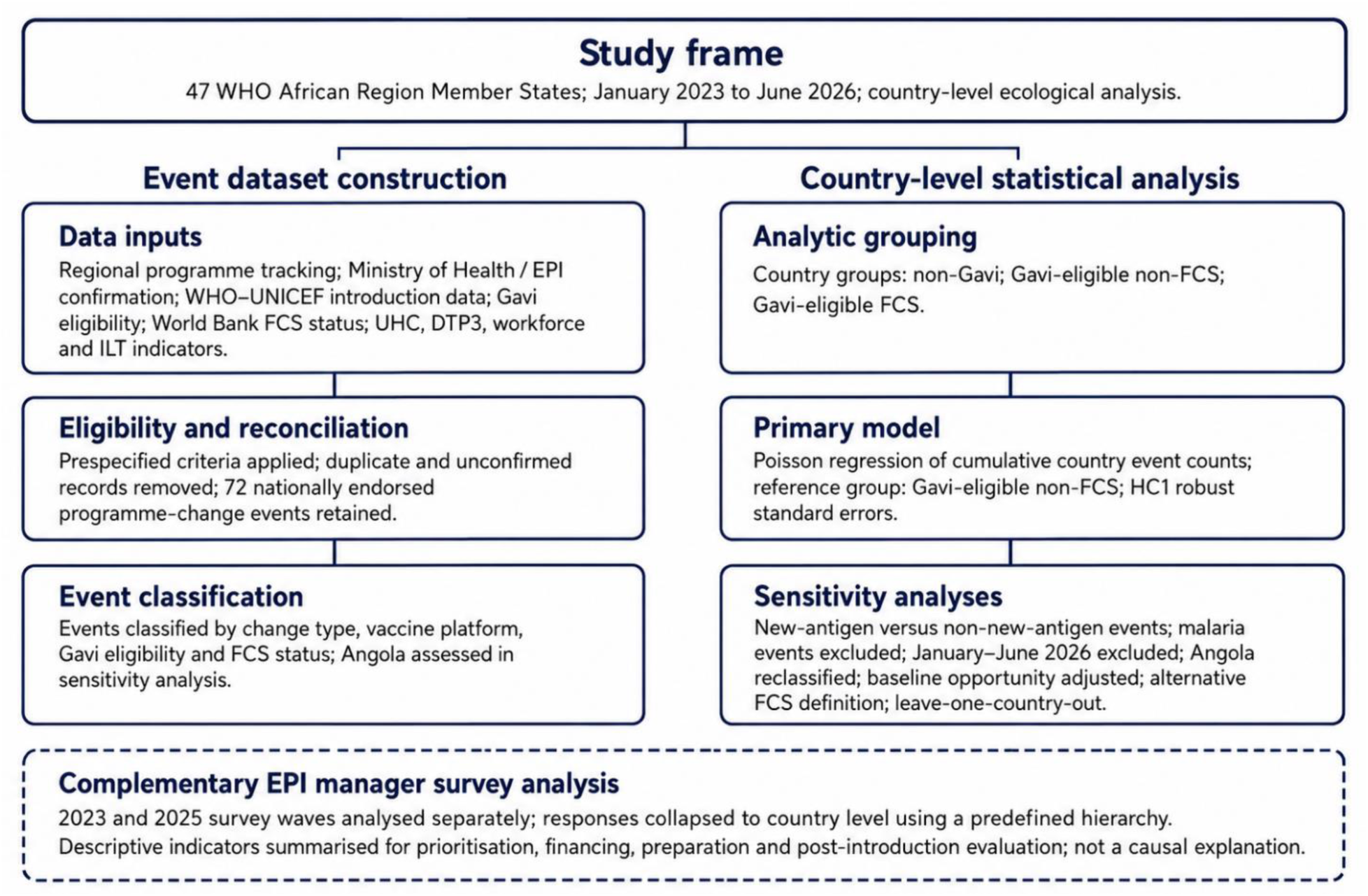
Study design and analysis flow. The figure summarises event dataset construction, country-level statistical analysis, sensitivity analyses, and the separate use of EPI manager survey data for programme contextualisation. EPI = Expanded Programme on Immunization; FCS = fragile and conflict-affected situations. Conflict-affected status refers to the World Bank FY26 conflict-affected category; broader FCS was examined only in sensitivity analysis; DTP3 = third-dose diphtheria–tetanus–pertussis coverage; ILT = Immunization League Table; UHC = universal health coverage service coverage index. This figure describes the analytic workflow, not a causal pathway.

### 2.2 Definition and classification of vaccine-programme-change events

A vaccine-programme-change event was defined as a nationally endorsed programme change requiring adaptation in planning, financing, procurement, delivery, recording and reporting, safety monitoring or post-introduction follow-up. Eligible events followed WHO guidance and included new disease-antigen introductions, new combination vaccines, and selected product, presentation, formulation, schedule or route-of-administration changes requiring programme adaptation [6,18]. Geographically targeted implementation was included only when it formed part of a national policy or programme plan. COVID-19 vaccine deployment, campaigns without routine-programme adoption, purely administrative changes and survey-only records not confirmed in regional tracking were excluded. Events were classified as new-antigen introductions, dose or schedule expansions, or combination-vaccine introductions. First national use of a newly WHO-prequalified and WHO-recommended vaccine through outbreak response was counted only when it required national endorsement, first entry into national supply and recording systems, product-specific delivery and safety-monitoring adaptation, and a documented pathway to preventive or routine programme use; this rule applied to two pentavalent meningococcal conjugate vaccine (Men5CV) events and is detailed, with the full inclusion criteria and exclusions, in Supplementary Tables S1 and S2 [28–30].

### 2.3 Data sources, linkage and contextual indicators

Regional programme-tracking data captured country, ISO3 code, subregion, year or period, vaccine or platform, and event type. These data were linked to a country master file containing Gavi eligibility, World Bank conflict-affected classification, DTP3 coverage, the universal health coverage (UHC) service coverage index, nurse and midwife density, and the 2025 Immunization League Table (ILT) score and band [4,20,31,32]. The ILT was used as a concurrent programme-performance marker because it was designed by the WHO Regional Office for Africa as a benchmarking and prioritisation tool to identify immunity and equity gaps and target differentiated support; it incorporates DTP3 and zero-dose information and was not treated as an externally validated readiness index, with its components and limitations documented in Supplementary Table S7 [32]. Country identifiers, vaccine labels and event classifications were standardised and validated against regional tracking records and Ministry of Health or EPI confirmation where available. All 47 Member States had complete outcome, financing–conflict and concurrent system-performance data.

### 2.4 Financing and conflict classification

The primary explanatory grouping was a three-level financing–conflict classification: non-Gavi, Gavi-eligible non-conflict-affected, and Gavi-eligible conflict-affected. Gavi eligibility was assigned according to eligibility status during the event year; Angola was handled as a prespecified sensitivity case because its eligibility changed in 2026, and was classified as non-Gavi in the primary analysis because its recorded events occurred in 2025 [33,34]. For the primary analysis, conflict-affected status was assigned using the World Bank FY26 conflict-affected category among WHO African Region Member States. Countries listed only under the World Bank institutional and social fragility category were not included in the primary conflict-affected group; this choice is reported explicitly because the World Bank classification distinguishes conflict-affected situations from institutional and social fragility, and a classification sensitivity using the broader fragile and conflict-affected situation (FCS) definition, which includes both categories, is reported in Supplementary Table S5 [19,20]. Because conflict-affected status was nested within Gavi eligibility, the three-level grouping avoided interpreting the conflict-affected contrast as a simple Gavi versus non-Gavi comparison. DTP3 coverage, the UHC service coverage index, nurse and midwife density, and the ILT score and band were analysed as concurrent system-performance indicators [31,32].

### 2.5 EPI manager survey data

Two EPI manager survey waves were analysed separately as programme-reported implementation data. Structured multilingual instruments were administered in English, French and Portuguese through WHO Country Offices. The 2023 wave captured prioritisation processes, decision-making constraints and willingness to adopt multi-criteria decision analysis (MCDA); the 2025 wave captured financing, donor-related delays or changes, preparation, product or formulation changes, partner support and post-introduction evaluation. Respondents received an information statement before survey access and participation was voluntary; responses were analysed as anonymised aggregate programme data, and country-level submissions were validated by the respective National EPI Manager and Ministry of Health before inclusion. Survey items on prioritisation and MCDA were interpreted against WHO decision-support principles for vaccine prioritisation, including structured appraisal of disease burden, vaccine performance, safety, affordability, sustainability, programme capacity and opportunity cost [11,35]. Because the two waves used different instruments and overlapping but non-identical respondents, they were analysed separately and not pooled; country-collapsed estimates, derived using a predefined respondent hierarchy, were the primary survey analysis, with item domains, denominators and respondent-level analyses in Supplementary Table S6. The survey findings were used as complementary programme intelligence and were not treated as formal qualitative data or as causal explanatory evidence.

### 2.6 Statistical analysis

The primary outcome was the cumulative number of vaccine-programme-change events per Member State. Analyses were conducted at country level (N=47); country-years were not treated as independent observations. No population offset was used, because all countries shared the same observation window and the outcome was a count of national programme-change events rather than population exposure, doses delivered or health impact. Poisson regression modelled event counts by financing–conflict group, with Gavi-eligible non-conflict-affected countries as the reference, reporting incidence rate ratios (IRRs) with 95% confidence intervals (CIs) using HC1 robust standard errors. Model fit was assessed using Pearson χ²/df and deviance/df; negative-binomial regression was fitted as an overdispersion check. Group-mean confidence intervals were estimated by nonparametric bootstrap with 10,000 resamples, resampling countries within group; the random seed is provided in Supplementary File S2. Fully adjusted multivariable models were not used as the primary specification, because of the small country denominator and event count.

Sensitivity analyses restricted the outcome to new-antigen introductions and to non-new-antigen events, excluded malaria-vaccine events, excluded partial-year 2026 entries, reclassified Angola as Gavi-eligible, adjusted for baseline introduction opportunity, applied the broader World Bank FCS definition, and used leave-one-country-out analysis. Baseline introduction opportunity was defined as the number of seven selected recommended vaccines or schedule components not yet introduced nationally by the end of 2022 (pneumococcal conjugate vaccine, rotavirus vaccine, hepatitis B birth dose, IPV2, second dose of measles-containing vaccine, rubella-containing vaccine and HPV vaccine), using WHO/UNICEF national introduction-year data; this proxy excluded targeted products such as malaria, typhoid and meningococcal vaccines (Supplementary Table S4) [4]. Fisher’s exact tests examined multiple-introduction status across ILT bands, and Spearman correlations assessed associations between event counts and concurrent system-performance indicators. Given the ecological design, small denominator and multiple exploratory comparisons, p values were interpreted as descriptive compatibility measures rather than causal evidence, and associations were described as associations rather than as drivers or effects. Analyses used Python 3.12 and R 4.4; code and a reproducibility README are provided in Supplementary File S2 [36–38]. No multiplicity correction was applied; secondary subgroup, correlation and sensitivity analyses were interpreted as exploratory compatibility analyses rather than confirmatory hypothesis tests.

### 2.7 Ethics and governance

This study used aggregate programme-level administrative, planning and survey data generated during routine immunisation programme monitoring and technical-support activities. No individual-level clinical data, patient records, biological samples or personally identifiable information were accessed. Because the activity constituted programme monitoring and evaluation using aggregate operational data collected as part of routine work, no additional research ethics committee review was requested or obtained; the full ethics and governance statement is provided under Declarations.

## 3. Results

### 3.1 Scale, timing and composition of vaccine-programme-change events

Across the 47 Member States, 38 countries (80.9%) recorded at least one vaccine-programme-change event between January 2023 and June 2026, while nine (19.1%) recorded none. Seventy-two events were recorded in total: 11 in 2023, 28 in 2024, 26 in 2025 and seven in January–June 2026. The 2026 records covered only the first half of the year and were not annualised. Events clustered in 2024–2025, which accounted for 54 of 72 events (75.0%). By event type, 48 events (66.7%) were new-antigen introductions, 20 (27.8%) were dose or schedule expansions, and four (5.6%) were combination-vaccine introductions. Malaria vaccines were the largest vaccine group (21 events, 29.2%: 12 RTS,S/AS01 and nine R21/Matrix-M), followed by HPV vaccine and IPV2 with 12 events each (16.7%), hepatitis B birth dose with six (8.3%), and typhoid conjugate and hexavalent vaccines with four each (5.6%); the remainder comprised pneumococcal conjugate, rotavirus and rubella-containing vaccines (three each) and second-dose measles-containing vaccine and Men5CV (two each) (Table 1; Supplementary Table S2).

**Table 1.** Composition of 72 vaccine-programme-change events by type and vaccine or platform, WHO African Region, January 2023–June 2026.

| Characteristic | Events | % of 72 |
| --- | --- | --- |
| **Event type** |  |  |
| New-antigen introduction | 48 | 66.7 |
| Dose or schedule expansion | 20 | 27.8 |
| Combination-vaccine introduction | 4 | 5.6 |
| **Vaccine or platform** |  |  |
| Malaria vaccine (RTS,S/AS01 12; R21/Matrix-M 9) | 21 | 29.2 |
| HPV vaccine | 12 | 16.7 |
| IPV2 | 12 | 16.7 |
| Hepatitis B birth dose | 6 | 8.3 |
| Typhoid conjugate vaccine | 4 | 5.6 |
| Hexavalent vaccine | 4 | 5.6 |
| Pneumococcal conjugate vaccine | 3 | 4.2 |
| Rotavirus vaccine | 3 | 4.2 |
| Rubella-containing or measles–rubella vaccine | 3 | 4.2 |
| Second-dose measles-containing vaccine (MCV2) | 2 | 2.8 |
| Men5CV (multivalent meningococcal conjugate) | 2 | 2.8 |
| **Total** | **72** | **100.0** |

Percentages use the 72 recorded events as the denominator. Event-type and vaccine/platform categories are separate classifications of the same 72 events. HPV = human papillomavirus vaccine; IPV2 = second dose of inactivated poliovirus vaccine; Men5CV = pentavalent meningococcal A/C/W/Y/X conjugate vaccine.

### 3.2 Subregional distribution

Subregional distributions differed in portfolio composition rather than performance. West Africa had the largest new-antigen component (24 of 30 events), driven mainly by malaria vaccines and HPV vaccine; Central Africa showed a mixed malaria-vaccine and IPV2 profile; and Eastern and Southern Africa had the largest dose or schedule-expansion component, led by IPV2 and hepatitis B birth dose. These differences were interpreted descriptively, because subregions differed in financing eligibility, disease burden, conflict exposure and remaining introduction opportunity; full subregional counts and means are reported in Supplementary Table S2.

### 3.3 Programme-change activity by financing and conflict status

The main gradient emerged where Gavi eligibility intersected with conflict-affected status, rather than from Gavi eligibility alone. Non-Gavi countries recorded 14 events across 11 countries, and Gavi-eligible non-conflict-affected countries recorded 33 events across 26 countries, both averaging 1.27 events per country; Gavi-eligible conflict-affected countries recorded 25 events across 10 countries, averaging 2.50 events per country (Figure 2). In the three-level Poisson model, Gavi-eligible conflict-affected status was associated with higher event counts (IRR 1.97, 95% CI 1.38–2.81; p<0.001), whereas non-Gavi status was not (IRR 1.00, 95% CI 0.54–1.87; p=0.99) (Table 2). Diagnostics did not indicate overdispersion (Pearson χ²/df 0.75; deviance/df 0.91), and negative-binomial regression gave a concordant estimate; reclassifying Angola as Gavi-eligible left the estimate essentially unchanged (IRR 1.93, 95% CI 1.36–2.73). The highest mean event count was observed in the group with the lowest median concurrent system-performance indicators, which are concurrent and descriptive and do not measure readiness or establish temporal prediction (Table 2).

**Figure 2.**
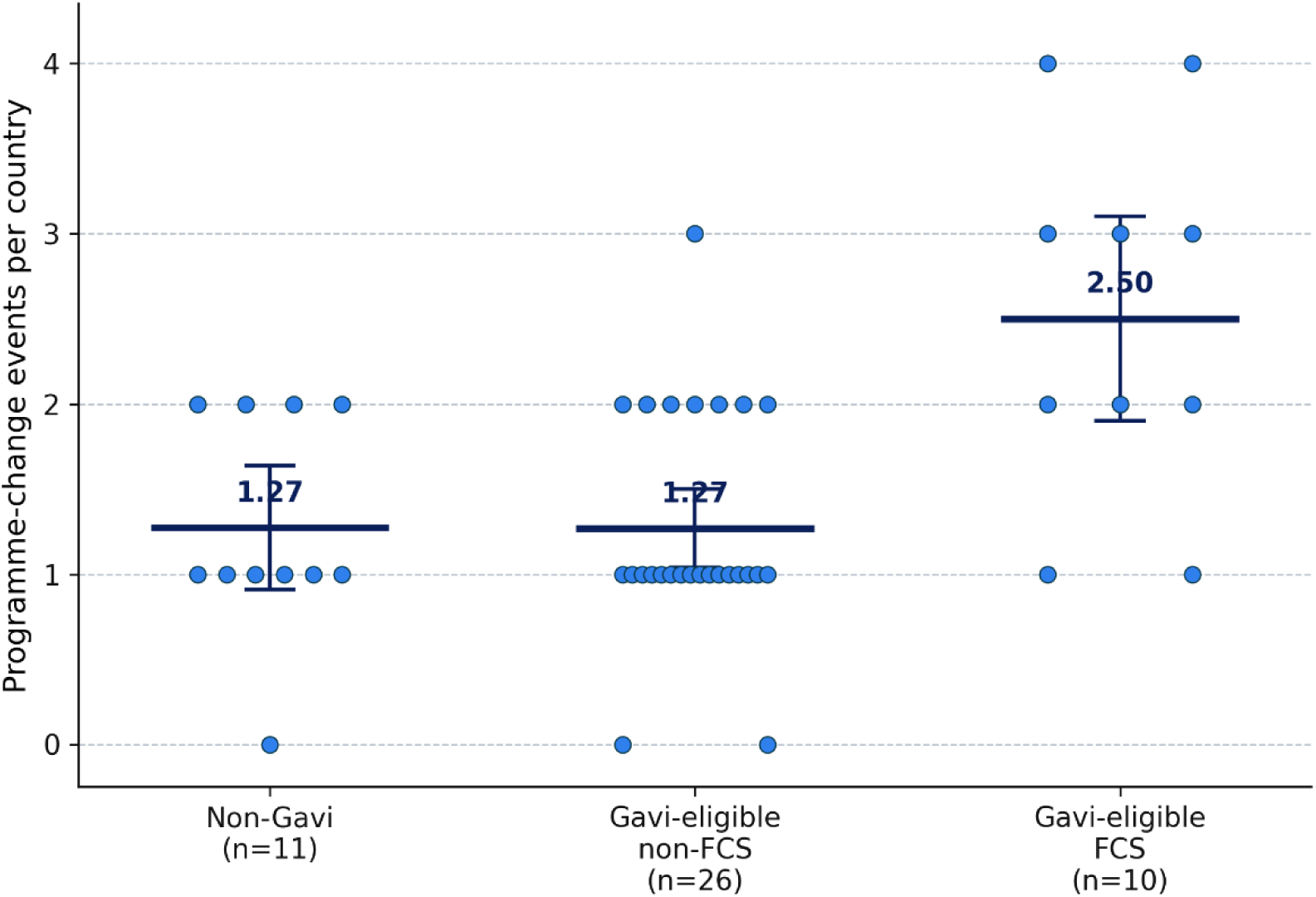
Programme-change events by financing–fragility group. Each point represents one country; points are horizontally jittered to reduce overlap. Horizontal bars show group means. The primary Poisson model compared Gavi-eligible FCS countries with Gavi-eligible non-FCS countries as the reference group and estimated an incidence rate ratio of 1.97, 95% confidence interval 1.38–2.81; p<0.001. FCS = fragile and conflict-affected situations; IRR = incidence rate ratio.

**Table 2.** Programme-change events and concurrent system-performance indicators by financing–conflict group (primary classification, Angola non-Gavi), WHO African Region, January 2023–June 2026.

| Group | Countries | Events | Mean/country (95% CI) | IRR (95% CI) | UHC index | DTP3 (%) | Workforce /1000 | ILT (%) |
| --- | --- | --- | --- | --- | --- | --- | --- | --- |
| Non-Gavi | 11 | 14 | 1.27 (0.55–2.00) | 1.00 (0.54–1.87) | 70.0 | 84.0 | 2.63 | 77.5 |
| Gavi-eligible, non-conflict-affected (ref.) | 26 | 33 | 1.27 (0.92–1.62) | 1.00 (ref.) | 48.5 | 86.5 | 0.79 | 62.5 |
| Gavi-eligible, conflict-affected | 10 | 25 | 2.50 (1.90–3.00) | 1.97 (1.38–2.81) | 41.0 | 73.0 | 0.45 | 48.8 |

### 3.4 New-antigen and non-new-antigen events

The financing–conflict gradient was concentrated in new-antigen introductions. Mean new-antigen events per country were 0.55 in non-Gavi countries, 0.85 in Gavi-eligible non-conflict-affected countries and 2.00 in Gavi-eligible conflict-affected countries; in the new-antigen model, Gavi-eligible conflict-affected status was associated with higher counts (IRR 2.36, 95% CI 1.43–3.91; p=0.001). By contrast, the association was not observed for non-new-antigen events (dose or schedule expansions and combination-vaccine introductions; conflict-affected IRR 1.13, 95% CI 0.37–3.41; p=0.84). The gradient was therefore concentrated in the event category with the greatest expected implementation complexity, including new delivery platforms, procurement decisions, safety monitoring and data-system adaptation.

### 3.5 Concurrent immunisation and health-system performance indicators

Higher programme-change activity co-occurred with weaker concurrent system-performance indicators. Countries in low or very-low ILT bands were more likely to record two or more events than countries in moderate or high bands (15 of 24, 62.5%, versus seven of 23, 30.4%; Fisher’s exact p=0.041); however, the ILT incorporates DTP3 and zero-dose information and was used only as a concurrent descriptive marker, so this association is partly mechanical and is not independent evidence. Event counts were also inversely associated with the UHC service coverage index; this and the other concurrent-indicator correlations are reported descriptively in Supplementary Table S7 and did not survive multiplicity adjustment. These analyses describe ecological co-occurrence, not temporal prediction, implementation readiness or causal effect, and because the ILT incorporates DTP3 and zero-dose information its association with event counts is partly mechanical and is reported descriptively rather than as an independent association.

### 3.6 EPI manager survey findings

Country-collapsed EPI manager survey findings identified implementation-assurance signals not captured by launch-status monitoring (Figure 3; Table 3). In the 2023 wave, 20 of 37 countries (54.1%) reported ad hoc vaccine-prioritisation processes, 19 of 37 (51.4%) identified limited resources or insufficient data as the main prioritisation challenge, and 30 of 37 (81.1%) expressed definite willingness to adopt a structured MCDA tool. In the 2025 wave, financing and sustainability was reported as a top implementation challenge by 28 of 35 countries (80.0%); donor-related delays or changes to introduction plans by seven of 35 (20.0%); and product or formulation changes over the preceding five years by 20 of 35 (57.1%). Among 25 countries reporting a planned 2026 introduction, preparatory activities had not started in nine (36.0%), and recent post-introduction evaluation was reported by 13 of 35 (37.1%). These country-reported indicators are descriptive, drawn from separate survey waves with item-specific denominators, and should not be interpreted as a longitudinal trend.

**Figure 3.**
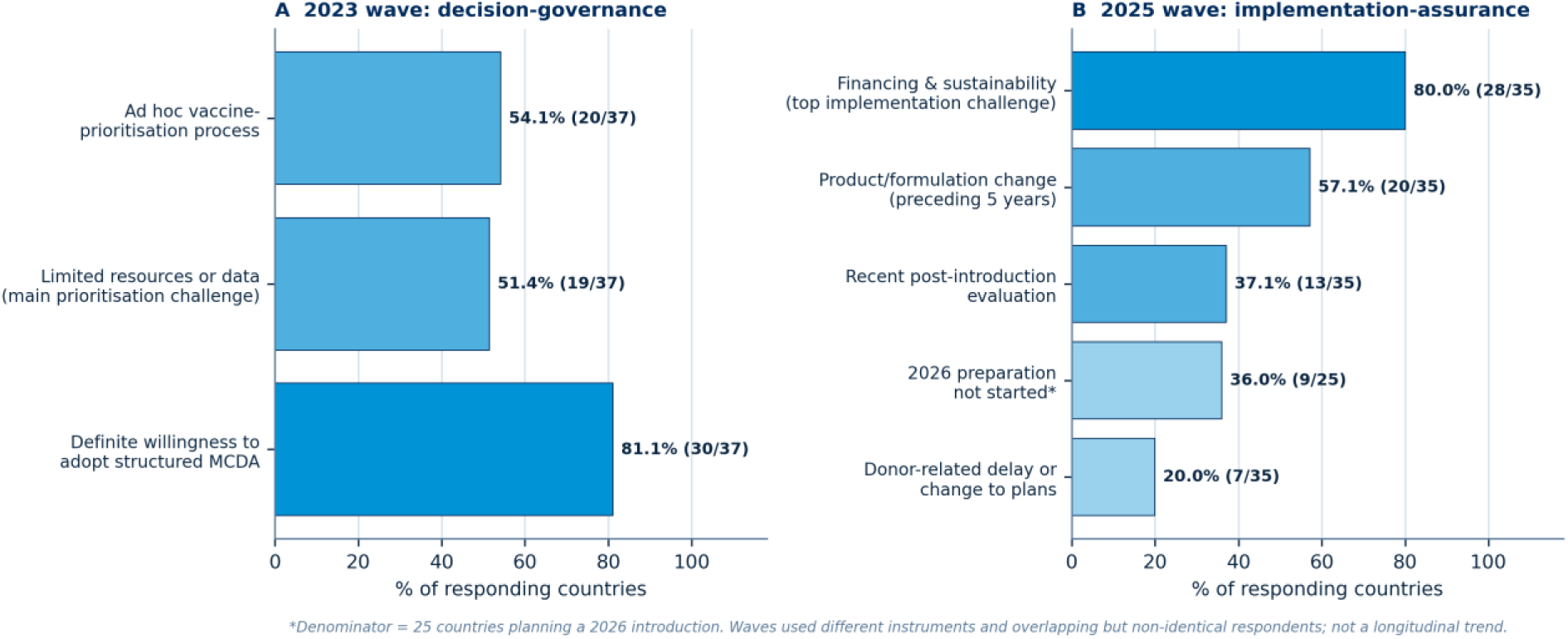
EPI manager-reported prioritisation, financing and implementation signals (country-collapsed), 2023 and 2025 waves. Bars show the percentage of responding countries reporting each constraint; the 2026-preparation denominator is the 25 countries planning a 2026 introduction. Composition of events by vaccine or platform and year, and the leave-one-country-out influence analysis, are provided as Supplementary Figures S1 and S2.

**Table 3.** Country-collapsed EPI manager survey indicators (primary analysis), 2023 and 2025 waves, WHO African Region.

| Indicator | Wave | n / N | % |
| --- | --- | --- | --- |
| Ad hoc prioritisation process | 2023 | 20 / 37 | 54.1 |
| Willing to adopt a structured MCDA tool | 2023 | 30 / 37 | 81.1 |
| Limited resources or data as main challenge | 2023 | 19 / 37 | 51.4 |
| Product or formulation switch, past 5 years | 2025 | 20 / 35 | 57.1 |
| Donor-related delay to introduction plans | 2025 | 7 / 35 | 20.0 |
| Preparatory activities not started (of planning countries) | 2025 | 9 / 25 | 36.0 |
| Recent post-introduction evaluation | 2025 | 13 / 35 | 37.1 |

### 3.7 Robustness analyses

The financing–conflict gradient was partly sensitive to malaria-vaccine activity, which accounted for 21 of 72 events and 10 of 25 events in the Gavi-eligible conflict-affected group. After excluding malaria-vaccine events, the association attenuated but remained in the same direction (Gavi-eligible conflict-affected mean 1.50 per country, versus 1.27 in non-Gavi and 0.85 in Gavi-eligible non-conflict-affected countries; conflict-affected IRR 1.77, 95% CI 1.06–2.97; p=0.030). The gradient also persisted after excluding January–June 2026 entries (IRR 2.24, 95% CI 1.51–3.33; p<0.001) and after adjustment for baseline introduction opportunity (IRR 2.00, 95% CI 1.36–2.94; p<0.001); remaining introduction opportunity itself was not associated with event counts (IRR 0.99, 95% CI 0.90–1.08; p=0.78). In leave-one-country-out analysis the conflict-affected IRR ranged from 1.84 to 2.10, indicating that no single country drove the primary estimate. The direction of the association was stable under the broader World Bank FCS definition, although attenuated: Gavi-eligible broader-FCS countries recorded 36 events across 18 countries, and the broader-FCS IRR was 1.64 (95% CI 1.08–2.47; p=0.019). Full numerical outputs are reported in Supplementary Table S5, and the leave-one-country-out analysis is reported in Supplementary Figure S2.

## 4. Discussion

This study reframes vaccine-introduction monitoring in the WHO African Region from a launch-counting exercise into a distributional implementation-risk question. The empirical signal is not that many events occurred, but that programme-change activity accumulated specifically at the Gavi–conflict interface, while non-Gavi and Gavi-eligible non-conflict-affected countries had similar event intensity. This matters because it was strongest for new-antigen introductions—the category most likely to require new delivery platforms, recurrent financing, safety monitoring and data-system adaptation. The signal should be read as an implementation-assurance concern, not as evidence that conflict-affected countries are introducing too many vaccines: many carry a high burden of vaccine-preventable disease, large zero-dose populations and a strong equity claim to accelerated access, so the policy issue is the co-location of epidemiological need, financing eligibility, product allocation and implementation risk, and the appropriate response is differentiated support rather than slower access.

Earlier regional studies established a historical adoption baseline rather than a distributional analysis: Sambala and colleagues documented major progress in new and underused vaccine adoption during the Decade of Vaccines and warned that transition, procurement, supply-chain capacity and sustainable domestic financing would determine whether gains could be sustained, and Iwu-Jaja and colleagues extended that baseline to 2022 [9,10]. Global monitoring shows that introduction of recommended vaccines remains incomplete and was slowed by the COVID-19 pandemic, with only 17% of countries having introduced all selected WHO-recommended new and underused vaccines by 2021 [3,39]. Recent timeliness evidence reinforces the distinction between adoption and protection: across 91 low- and middle-income countries, only about half of DTP3 and second-dose measles-containing vaccine doses were given within 28 days of the target age [17]. The IA2030 mid-term review reaches a similar operational diagnosis, calling for stronger country ownership, tailored support, subnational data use and focused support to fragile, conflict-affected and vulnerable settings; this study contributes regional evidence on where programme-change activity is accumulating [5].

The main gradient was not explained by Gavi eligibility alone: non-Gavi and Gavi-eligible non-conflict-affected countries had similar mean event counts, whereas Gavi-eligible conflict-affected countries had substantially higher activity, suggesting that recent activity is shaped by the intersection of financing eligibility, epidemiological need, product allocation, partner support and operational context rather than by eligibility status in isolation [33,34]. This interpretation is consistent with political-economy scholarship on global health initiatives, which cautions that programme outputs can obscure the country-level practices, competition over resources and accountability effects generated by financing and governance architectures [8]. The findings also point to a measurement asymmetry: launch is highly visible in global scorecards, while completion, continuity, safety, financing assurance and equity are less consistently measured. This is a structural feature of a monitoring architecture that has made access easier to count than sustained protection, and one that country vaccine budgets, portfolio optimisation, financing review and sequencing under Gavi 6.0 are intended to address [40]. The study did not measure financing flows, partner inputs, procurement prices or domestic budget execution, so this is an evidence-informed interpretation, not a causal claim. The system-performance indicators were measured contemporaneously with the observation window, so the analysis cannot establish whether weaker systems preceded programme-change activity, followed it, or co-evolved with it under shared financing, conflict and delivery conditions.

The equity interpretation is central but must be framed precisely. This analysis did not measure subnational coverage, timeliness, dose completion, stock continuity, safety reporting, gender equity, displacement status or zero-dose reach, and therefore cannot show whether recent activity reduced or widened inequities. What it shows is an equity-risk configuration: programme-change activity accumulated where concurrent system-performance indicators were weaker and recurrent functions are harder to sustain, creating an inverse-care risk in which populations with the greatest expected marginal benefit may also be most likely to experience delayed, incomplete or interrupted vaccination. This is consistent with evidence that conflict-affected and broader fragile settings account for a disproportionate share of zero-dose children [3,5]. Different products expose different delivery dependencies: malaria vaccines require immunisation–malaria coordination, readiness monitoring and multi-dose completion, with documented dose attrition and financing exposure [13,41]; HPV vaccination requires adolescent, school and community platforms and remains far below the 90% target, with persistent gaps for out-of-school girls [15]; and timeliness evidence shows that delayed vaccination leaves children unprotected even when eventual coverage improves [17]. The issue is not whether these countries should introduce vaccines, but whether launch is accompanied by the delivery and equity functions required to reach populations already least served by routine systems [43,44].

Introduction decisions distribute responsibility across National Immunization Technical Advisory Groups (NITAGs), ministries of health and finance, procurement systems, regulators, pharmacovigilance authorities, subnational managers and partners; this accountability chain is meaningful only if linked to measurable post-launch functions—financing assurance, supply continuity, health-worker preparation, safety monitoring, demand generation, data-system adaptation, dose completion and post-introduction evaluation [7,34]. Structured prioritisation frameworks are useful less for producing a numerical ranking than for forcing decision-makers to make explicit the criteria, evidence gaps, opportunity costs and capacity requirements attached to each decision [11,22,35]. The EPI manager surveys reinforce this, identifying constraints in ad hoc prioritisation, limited resources or data, donor-related delays, product or formulation changes, delayed preparation and limited recent post-introduction evaluation; these signals indicate where regional and partner support can be better differentiated, sequenced and institutionalised rather than country failure [45].

The findings should not portray African countries as passive recipients of externally financed products. Countries exercise agency through prioritisation, advisory deliberation, ministry negotiation, surveillance interpretation, regulatory decisions and delivery-platform design, but that agency operates within financing rules, product availability, market conditions and partner timelines [8,46]. Consistent with recent regional strategic analysis, the next decade will require fiscal and policy stewardship, structured health–finance–planning dialogue, consolidation of fragmented platforms, regional regulatory capacity, local evidence generation and investment in African vaccine manufacturing and supply resilience, as the model of externally financed, disease-specific programming weakens [46,47]. The IA2030 mid-term review similarly places countries and regions at the centre of strategic decision-making [5].

### Strengths and limitations

This study covers all 47 Member States, uses country-level analysis rather than country-year pseudo-replication, distinguishes new-antigen introductions from dose or schedule expansions and combination-vaccine introductions, links event counts to financing–conflict and concurrent system-performance indicators, and triangulates tracking data with two EPI manager survey waves. Several limitations define the boundary of inference. First, the outcome was an unweighted country-level event count, not a measure of doses delivered, target-population size, recurrent cost, coverage, dose completion, stock continuity, safety reporting, equity or health impact, and events differed substantially in operational footprint. Second, adjustment for baseline introduction opportunity reduced, but did not eliminate, the concern that the gradient reflected differential room to introduce, and the opportunity proxy excluded targeted products such as malaria, typhoid and meningococcal vaccines. Third, the gradient was sensitive to malaria-vaccine roll-out and was materially attenuated when malaria events were excluded (conflict-affected IRR 1.77, with a confidence interval approaching unity), so the headline estimate should be read as a pattern observed during a period of major malaria-vaccine expansion rather than a stable structural law, although leave-one-country-out analysis (IRR 1.84–2.10) indicated that no single country drove it. Fourth, the primary exposure used the World Bank conflict-affected category among Member States; countries listed under institutional and social fragility but not conflict were not included in the primary group, which improves interpretability of the conflict-related operational context but means the findings should not be generalised to all forms of institutional or social fragility. The broader-FCS sensitivity attenuated the estimate, supporting the interpretation that the primary signal was strongest for conflict-affected settings rather than for all forms of institutional or social fragility. Fifth, the design is ecological and descriptive and supports no causal inference; with 47 countries the models were sparse, the primary contrast rested on 10 conflict-affected countries and 25 events, and the correlational analyses were unadjusted for multiplicity and are hypothesis-generating. Sixth, the system-performance indicators are concurrent and partly non-independent, because the ILT incorporates DTP3 and zero-dose information. Seventh, the surveys were self-reported institutional submissions returned by 37 of 47 (2023) and 35 of 47 (2025) countries, so non-response may bias these descriptive estimates, which should be read as programme intelligence rather than prevalence. Because first national use of a newly recommended product through outbreak response can blur the boundary between campaign and programme change, the Men5CV classification rule was prespecified and documented, and the two Men5CV events could not materially influence the primary estimates. Finally, as a reflexive consideration, most authors are WHO staff who support the introductions analysed; this positionality was addressed through predefined classifications, transparent denominators, reproducible analyses and explicit limitations, and the interpretation deliberately avoids locating responsibility only at country level [48].

### 4.8 Policy implications

The policy implication is differentiated implementation assurance, not delayed access; the indicators proposed here are recommended monitoring functions rather than outcomes measured in this study. For Gavi-eligible conflict-affected countries, assurance should begin at approval and include financing review, security-adapted microplanning, stock monitoring, safety surveillance and subnational equity review. For non-Gavi and transitioning countries, whose event intensity matched that of Gavi-eligible non-conflict-affected countries but whose financing regime differs, support should focus on procurement intelligence, price benchmarking, regulatory reliance and domestic-financing strategy [40,49]. Product-differentiated monitoring should be illustrative rather than a universal checklist: dose completion and stock continuity for multi-dose products [41]; school and out-of-school reach and district-level equity for adolescent vaccines [15]; timeliness for birth-dose and infant-schedule products [17]; and procurement lead time, cold-chain effects, wastage, training completion and safety reporting for switches and combination vaccines. Integrated delivery may reduce fragmentation under fiscal pressure, but effectiveness and equity should take precedence over short-term cost savings [45].

## 5. Conclusion

Between January 2023 and June 2026, vaccine-programme-change activity in the WHO African Region concentrated at the intersection of Gavi eligibility, conflict-affected status and weaker concurrent system-performance indicators, with the strongest gradient observed for new-antigen introductions. The association attenuated after excluding malaria-vaccine events and should not be interpreted causally. The policy implication is not to slow access in high-need settings, but to govern introduction as a multi-year commitment: financing review before approval, differentiated implementation support for conflict-affected settings, explicit inclusion of non-Gavi and transitioning countries, and post-launch accountability for coverage, dose completion, safety, stock continuity and subnational equity. Introduction should be the beginning of responsibility, not the end of measurement.

## Supporting information

STROBE RECORD checklist

## Data Availability

The de-identified analytic dataset, country master file, data dictionary and analysis code will be deposited in a public repository on journal acceptance, subject to institutional clearance. Public source indicators are available from the cited WHO, UNICEF, World Bank and Gavi sources. The Immunization League Table is an internal WHO programme document and is not publicly available; derived, non-identifiable analytic variables are provided where institutional clearance permits.

## Declarations

### Ethics approval and consent

This study used aggregate programme-level administrative, planning and survey data generated during routine immunisation programme monitoring and technical-support activities in the WHO African Region. No individual-level clinical data, patient records, biological samples or personally identifiable information were accessed. The EPI manager surveys were institutional programme-planning and monitoring instruments; respondents received information on the purpose of the survey before participation, participation was voluntary, and responses were analysed only in anonymised, aggregate or country-collapsed form. Country-level submissions used in the analysis were validated by National EPI Managers or Ministries of Health before inclusion. Because the activity constituted programme monitoring and evaluation using aggregate operational data collected as part of routine work, no additional research ethics committee review was requested or obtained.

## Acknowledgements

The authors thank EPI managers and Ministry of Health colleagues in WHO African Region Member States who provided and validated country data, and WHO IST and Country Office immunisation focal points who supported survey distribution.

## CRediT author contributions

Adidja Amani: Conceptualization, Methodology, Investigation, Data curation, Formal analysis, Writing – original draft, Writing – review and editing, Supervision, Project administration. Pamela Mitula: Investigation, Data curation, Validation, Writing – review and editing. Celestin Danwang: Methodology, Validation, Writing – review and editing. Aschalew Teka Bekele: Investigation, Validation, Writing – review and editing. Hermann Didi Ngossaki: Validation, Writing – review and editing. Yolande Vuo Masembe: Investigation, Validation, Writing – review and editing. Lanzy Achille: Writing – review and editing. Claude Mangobo: Validation, Writing – review and editing. Ado Mpia Bwaka: Investigation, Validation, Writing – review and editing. Marcellin Nimpa Mengouo: Investigation, Validation, Writing – review and editing. Mutale Mumba: Investigation, Validation, Writing – review and editing. Terna Nomhwange: Investigation, Validation, Writing – review and editing. Amavi Edinam Agbenu: Investigation, Validation, Writing – review and editing. Nosheen Safdar: Writing – review and editing. André Arsène Bita Fouda: Writing – review and editing. Alam Khattak: Conceptualization, Interpretation, Writing – review and editing. Lynda Rey: Writing – review and editing. Okot Charles Lukoya: Investigation, Validation, Writing – review and editing. Kwasi Nyarko: Investigation, Validation, Writing – review and editing. Bridget Lindsay Farham: Writing – review and editing. Anuj Kapilashrami: Conceptualization, Supervision, Writing – review and editing. Akpaka A. Kalu: Supervision, Interpretation, Writing – review and editing. Benido Impouma: Supervision, Conceptualization, Interpretation, Writing – review and editing. All authors approved the final manuscript and agreed to be accountable for all aspects of the work.

## Declaration of generative AI in the writing process

During the preparation of this work, the authors used AI to support language editing and improve readability. After using these tools, the authors reviewed and edited the content as needed and take full responsibility for the content of the published article. AI tools were not used to generate data, conduct the statistical analysis, or produce scientific conclusions, and are not listed as authors.

