## Supplementary Material for "Vaccine introductions in the WHO African Region, 2023–26: a country-level ecological analysis by Gavi eligibility and conflict-affected status"

This appendix contains Supplementary Tables S1–S7, Supplementary Figures S1–S2, and Supplementary File descriptions S1–S2. All tables reconcile to 72 nationally endorsed vaccine-programme-change events across the 47 WHO African Region Member States, of which 38 recorded at least one event. Numerals use the full point as the decimal separator. The complete machine-readable event-level dataset, the 47-country master file with all per-country indicators, the baseline introduction-opportunity index and the analysis code will be deposited in a public repository on acceptance, subject to institutional clearance. High-resolution figure files are available for journal submission and revision.

### Contents


**Table S1** Eligibility, inclusion and exclusion rules for vaccine-programme-change events

**Table S2** Analytic event inventory by vaccine or platform and event type, with the full 72-event enumeration

**Table S3** Country master file and financing–conflict classification (47 Member States)

**Table S4** Baseline introduction-opportunity index

**Table S5** Primary and sensitivity model outputs

**Table S6** EPI manager survey indicators, domains and denominators

**Table S7** Immunization League Table (ILT): composition, interpretation and concurrent-indicator correlations

**Figure S1** Event composition by period and vaccine or platform

**Figure S2** Robustness and leave-one-country-out influence analysis

**File S1** STROBE and RECORD reporting

**File S2** Reproducibility and analysis code

### Supplementary Table S1. Eligibility, inclusion and exclusion rules for vaccine-programme-change events

A vaccine-programme-change event was a nationally endorsed programme change requiring adaptation in planning, financing, procurement, delivery, recording and reporting, safety monitoring or post-introduction follow-up. The rules below were applied to construct the final 72-event analytic dataset from regional programme-tracking data, Ministry of Health or EPI confirmation, WHO/UNICEF introduction data and product-specific documentation.

| Rule | Record type | Inclusion criterion | Exclusion / rationale |
|---|---|---|---|
| R1 | New-antigen introduction | Nationally endorsed first introduction of a vaccine against a new disease antigen into the national programme | Subnational pilots without national policy |
| R2 | Dose or schedule expansion | Nationally endorsed added dose or schedule change for an existing antigen | Administrative recording-only updates |
| R3 | Combination-vaccine introduction | Switch to, or addition of, a combination product that changed the routine programme | Same product repackaged only |
| R4 | Product / presentation / formulation / route change | Programme-relevant change requiring procurement, cold-chain, training, safety or data-system adaptation | Like-for-like restock |
| R5 | Geographically targeted introduction | Targeted use forming part of a national policy or programme plan | Ad hoc local activity without national endorsement |
| R6–R8 | Campaign without routine adoption; COVID-19 deployment; purely administrative change | Not counted as events | Outside the programme-change definition; emergency/separately financed response |
| R9 | Survey-only record | Counted only if confirmed in regional tracking | Excluded if unconfirmed (avoids double counting) |
| R10 | Men5CV first national use via outbreak response | Counted only with national endorsement, first entry into national supply and recording systems, product-specific delivery and safety-monitoring adaptation, and a documented pathway to preventive or routine use | Reactive use of established products without programme change |

*Rule R10 applied to the two pentavalent meningococcal conjugate vaccine (Men5CV) events (Nigeria 2024; Niger 2025) and keeps outbreak-response activity analytically distinct from routine programme introduction.*

### Supplementary Table S2. Analytic event inventory by vaccine or platform and event type

The verified aggregate inventory reconciles to 72 events and is the companion to main-text Table 1. Panel A cross-classifies the 72 events by vaccine or platform and event type; Panel B summarises the distribution by period, financing–conflict group and subregion; Panel C lists every included event.

#### Panel A. Events by vaccine or platform × event type

| Vaccine / platform | New-antigen | Dose / schedule | Combination | Total |
|---|---|---|---|---|
| Malaria vaccine (RTS,S/AS01 12; R21/Matrix-M 9) | 21 | 0 | 0 | 21 |
| HPV vaccine | 12 | 0 | 0 | 12 |
| IPV2 | 0 | 12 | 0 | 12 |
| Hepatitis B birth dose | 6 | 0 | 0 | 6 |
| Typhoid conjugate vaccine (TCV) | 4 | 0 | 0 | 4 |
| Hexavalent vaccine | 0 | 0 | 4 | 4 |
| Pneumococcal conjugate vaccine (PCV) | 1 | 2 | 0 | 3 |
| Rotavirus vaccine | 1 | 2 | 0 | 3 |
| Rubella-containing / measles–rubella vaccine | 1 | 2 | 0 | 3 |
| Second-dose measles-containing vaccine (MCV2) | 0 | 2 | 0 | 2 |
| Men5CV (pentavalent meningococcal conjugate) | 2 | 0 | 0 | 2 |
| **Total** | **48** | **20** | **4** | **72** |

#### Panel B. Distribution by period, financing–conflict group and subregion

| Period | Events | % of 72 |
|---|---|---|
| 2023 | 11 | 15.3 |
| 2024 | 28 | 38.9 |
| 2025 | 26 | 36.1 |
| January–June 2026 | 7 | 9.7 |
| **Total** | **72** | **100.0** |

| Financing–conflict group | Events | Countries | Mean / country |
|---|---|---|---|
| Non-Gavi | 14 | 11 | 1.27 |
| Gavi-eligible, non-conflict-affected | 33 | 26 | 1.27 |
| Gavi-eligible, conflict-affected | 25 | 10 | 2.50 |
| **Total** | **72** | **47** | **1.53** |

| Subregion | Events | Countries | Active* | Mean / country | Mean / active |
|---|---|---|---|---|---|
| West Africa | 30 | 17 | 15 | 1.76 | 2.00 |
| Central Africa | 15 | 10 | 7 | 1.50 | 2.14 |
| Eastern and Southern Africa | 27 | 20 | 16 | 1.35 | 1.69 |
| **Total** | **72** | **47** | **38** | **1.53** | **1.89** |

**Active = countries with at least one included event. Subregions follow the WHO African Region programme structure and are descriptive, not performance rankings.*

### Panel C. Full 72-event analytic enumeration

All 72 included events are listed below in chronological order. The event identifier is an analytic identifier only and is not a source-record identifier. Subregion for each country is given in Supplementary Table S3.

| ID | Country | ISO3 | Period | Vaccine / platform | Event type | Financing–conflict group |
|---|---|---|---|---|---|---|
| E001 | Cabo Verde | CPV | 2023 | IPV2 | Dose or schedule expansion | Non-Gavi |
| E002 | Cameroon | CMR | 2023 | IPV2 | Dose or schedule expansion | Gavi, conflict-affected |
| E003 | Côte d’Ivoire | CIV | 2023 | IPV2 | Dose or schedule expansion | Gavi, non-conflict-affected |
| E004 | Dem. Rep. of the Congo | COD | 2023 | IPV2 | Dose or schedule expansion | Gavi, conflict-affected |
| E005 | Eswatini | SWZ | 2023 | HPV | New-antigen introduction | Non-Gavi |
| E006 | Malawi | MWI | 2023 | TCV | New-antigen introduction | Gavi, non-conflict-affected |
| E007 | Nigeria | NGA | 2023 | HPV | New-antigen introduction | Gavi, conflict-affected |
| E008 | Seychelles | SYC | 2023 | HepB birth dose | Dose or schedule expansion | Non-Gavi |
| E009 | South Africa | ZAF | 2023 | HepB birth dose | Dose or schedule expansion | Non-Gavi |
| E010 | Togo | TGO | 2023 | HPV | New-antigen introduction | Gavi, non-conflict-affected |
| E011 | Zambia | ZMB | 2023 | HPV | New-antigen introduction | Gavi, non-conflict-affected |
| E012 | Benin | BEN | 2024 | MCV2 | Dose or schedule expansion | Gavi, non-conflict-affected |
| E013 | Benin | BEN | 2024 | RTS,S/AS01 | New-antigen introduction | Gavi, non-conflict-affected |
| E014 | Botswana | BWA | 2024 | IPV2 | Dose or schedule expansion | Non-Gavi |
| E015 | Burkina Faso | BFA | 2024 | RTS,S/AS01 | New-antigen introduction | Gavi, conflict-affected |
| E016 | Cameroon | CMR | 2024 | RTS,S/AS01 | New-antigen introduction | Gavi, conflict-affected |
| E017 | Central African Republic | CAF | 2024 | RTS,S/AS01 | New-antigen introduction | Gavi, conflict-affected |
| E018 | Chad | TCD | 2024 | PCV | New-antigen introduction | Gavi, non-conflict-affected |
| E019 | Chad | TCD | 2024 | RTS,S/AS01 | New-antigen introduction | Gavi, non-conflict-affected |
| E020 | Chad | TCD | 2024 | Rotavirus | New-antigen introduction | Gavi, non-conflict-affected |
| E021 | Comoros | COM | 2024 | IPV2 | Dose or schedule expansion | Gavi, non-conflict-affected |
| E022 | Congo | COG | 2024 | IPV2 | Dose or schedule expansion | Gavi, non-conflict-affected |
| E023 | Côte d’Ivoire | CIV | 2024 | R21/Matrix-M | New-antigen introduction | Gavi, non-conflict-affected |
| E024 | Dem. Rep. of the Congo | COD | 2024 | R21/Matrix-M | New-antigen introduction | Gavi, conflict-affected |
| E025 | Eswatini | SWZ | 2024 | IPV2 | Dose or schedule expansion | Non-Gavi |

| ID | Country | ISO3 | Period | Vaccine / platform | Event type | Financing–conflict group |
|---|---|---|---|---|---|---|
| E026 | Ethiopia | ETH | 2024 | IPV2 | Dose or schedule expansion | Gavi, conflict-affected |
| E027 | Liberia | LBR | 2024 | RTS,S/AS01 | New-antigen introduction | Gavi, non-conflict-affected |
| E028 | Malawi | MWI | 2024 | IPV2 | Dose or schedule expansion | Gavi, non-conflict-affected |
| E029 | Mali | MLI | 2024 | HPV | New-antigen introduction | Gavi, conflict-affected |
| E030 | Mali | MLI | 2024 | Rubella-containing (RCV) | New-antigen introduction | Gavi, conflict-affected |
| E031 | Mozambique | MOZ | 2024 | R21/Matrix-M | New-antigen introduction | Gavi, conflict-affected |
| E032 | Niger | NER | 2024 | RTS,S/AS01 | New-antigen introduction | Gavi, conflict-affected |
| E033 | Nigeria | NGA | 2024 | Men5CV | New-antigen introduction | Gavi, conflict-affected |
| E034 | Nigeria | NGA | 2024 | R21/Matrix-M | New-antigen introduction | Gavi, conflict-affected |
| E035 | Seychelles | SYC | 2024 | Hexavalent | Combination-vaccine introduction | Non-Gavi |
| E036 | Sierra Leone | SLE | 2024 | RTS,S/AS01 | New-antigen introduction | Gavi, non-conflict-affected |
| E037 | South Africa | ZAF | 2024 | Rubella-containing (RCV) | New-antigen introduction | Non-Gavi |
| E038 | South Sudan | SSD | 2024 | RTS,S/AS01 | New-antigen introduction | Gavi, conflict-affected |
| E039 | Zambia | ZMB | 2024 | IPV2 | Dose or schedule expansion | Gavi, non-conflict-affected |
| E040 | Angola | AGO | 2025 | HPV | New-antigen introduction | Non-Gavi |
| E041 | Angola | AGO | 2025 | IPV2 | Dose or schedule expansion | Non-Gavi |
| E042 | Benin | BEN | 2025 | HPV | New-antigen introduction | Gavi, non-conflict-affected |
| E043 | Burkina Faso | BFA | 2025 | TCV | New-antigen introduction | Gavi, conflict-affected |
| E044 | Burundi | BDI | 2025 | RTS,S/AS01 | New-antigen introduction | Gavi, non-conflict-affected |
| E045 | Comoros | COM | 2025 | HPV | New-antigen introduction | Gavi, non-conflict-affected |
| E046 | Dem. Rep. of the Congo | COD | 2025 | MCV2 | Dose or schedule expansion | Gavi, conflict-affected |
| E047 | Eritrea | ERI | 2025 | HepB birth dose | Dose or schedule expansion | Gavi, non-conflict-affected |
| E048 | Ethiopia | ETH | 2025 | HepB birth dose | Dose or schedule expansion | Gavi, conflict-affected |
| E049 | Ethiopia | ETH | 2025 | R21/Matrix-M | New-antigen introduction | Gavi, conflict-affected |
| E050 | Ghana | GHA | 2025 | HPV | New-antigen introduction | Gavi, non-conflict-affected |
| E051 | Guinea | GIN | 2025 | RTS,S/AS01 | New-antigen introduction | Gavi, non-conflict-affected |
| E052 | Kenya | KEN | 2025 | TCV | New-antigen introduction | Gavi, non-conflict-affected |
| E053 | Madagascar | MDG | 2025 | HPV | New-antigen introduction | Gavi, non-conflict-affected |
| E054 | Mali | MLI | 2025 | R21/Matrix-M | New-antigen introduction | Gavi, conflict-affected |
| E055 | Mauritania | MRT | 2025 | Hexavalent | Combination-vaccine introduction | Gavi, non-conflict-affected |

| ID | Country | ISO3 | Period | Vaccine / platform | Event type | Financing–conflict group |
|---|---|---|---|---|---|---|
| E056 | Namibia | NAM | 2025 | HPV | New-antigen introduction | Non-Gavi |
| E057 | Niger | NER | 2025 | Men5CV | New-antigen introduction | Gavi, conflict-affected |
| E058 | Niger | NER | 2025 | TCV | New-antigen introduction | Gavi, conflict-affected |
| E059 | Nigeria | NGA | 2025 | Measles–rubella (MR) | New-antigen introduction | Gavi, conflict-affected |
| E060 | Senegal | SEN | 2025 | Hexavalent | Combination-vaccine introduction | Gavi, non-conflict-affected |
| E061 | South Sudan | SSD | 2025 | PCV | New-antigen introduction | Gavi, conflict-affected |
| E062 | South Sudan | SSD | 2025 | Rotavirus | New-antigen introduction | Gavi, conflict-affected |
| E063 | Togo | TGO | 2025 | R21/Matrix-M | New-antigen introduction | Gavi, non-conflict-affected |
| E064 | Uganda | UGA | 2025 | R21/Matrix-M | New-antigen introduction | Gavi, non-conflict-affected |
| E065 | Zambia | ZMB | 2025 | R21/Matrix-M | New-antigen introduction | Gavi, non-conflict-affected |
| E066 | Burundi | BDI | 2026 H1 | HPV | New-antigen introduction | Gavi, non-conflict-affected |
| E067 | Cabo Verde | CPV | 2026 H1 | Hexavalent | Combination-vaccine introduction | Non-Gavi |
| E068 | Cabo Verde | CPV | 2026 H1 | PCV | New-antigen introduction | Non-Gavi |
| E069 | Cabo Verde | CPV | 2026 H1 | Rotavirus | New-antigen introduction | Non-Gavi |
| E070 | Congo | COG | 2026 H1 | HepB birth dose | Dose or schedule expansion | Gavi, non-conflict-affected |
| E071 | Guinea-Bissau | GNB | 2026 H1 | RTS,S/AS01 | New-antigen introduction | Gavi, non-conflict-affected |
| E072 | Rwanda | RWA | 2026 H1 | HepB birth dose | Dose or schedule expansion | Gavi, non-conflict-affected |

*Reconciliation: 72 events; 48 new-antigen introductions, 20 dose or schedule expansions, 4 combination-vaccine introductions. Men5CV labels the two pentavalent meningococcal conjugate vaccine events. Group labels use the manuscript primary classification (World Bank FY26 conflict-affected category); the broader fragile and conflict-affected situation [FCS] grouping is reported only in Supplementary Table S5.*

### Supplementary Table S3. Country master file and financing–conflict classification (47 Member States)

One row per Member State. The table reconciles to 47 countries and 72 events. Per-country DTP3 coverage, the UHC service coverage index, nurse and midwife density and the 2025 Immunization League Table score and band will be provided with the deposited country master file; group medians appear in main-text Table 2.

| Country | ISO3 | Subregion | Gavi | Financing–conflict group | Confl. | Frag. | Br.-FCS | Events |
|---|---|---|---|---|---|---|---|---|
| Algeria | DZA | West | Non-Gavi | Non-Gavi | No | No | No | 0 |
| Angola* | AGO | Central | Non-Gavi | Non-Gavi | No | No | No | **2** |
| Benin | BEN | West | Gavi | Gavi, non-conflict-affected | No | No | No | **3** |
| Botswana | BWA | E. & Southern | Non-Gavi | Non-Gavi | No | No | No | **1** |
| Burkina Faso | BFA | West | Gavi | Gavi, conflict-affected | Yes | No | Yes | **2** |
| Burundi | BDI | Central | Gavi | Gavi, non-conflict-affected | No | Yes | Yes | **2** |
| Cabo Verde | CPV | West | Non-Gavi | Non-Gavi | No | No | No | **4** |
| Cameroon | CMR | Central | Gavi | Gavi, conflict-affected | Yes | No | Yes | **2** |
| Central African Republic | CAF | Central | Gavi | Gavi, conflict-affected | Yes | No | Yes | **1** |
| Chad | TCD | Central | Gavi | Gavi, non-conflict-affected | No | Yes | Yes | **3** |
| Comoros | COM | E. & Southern | Gavi | Gavi, non-conflict-affected | No | Yes | Yes | **2** |
| Congo | COG | Central | Gavi | Gavi, non-conflict-affected | No | Yes | Yes | **2** |
| Côte d’Ivoire | CIV | West | Gavi | Gavi, non-conflict-affected | No | No | No | **2** |
| Democratic Republic of the Congo | COD | Central | Gavi | Gavi, conflict-affected | Yes | No | Yes | **3** |
| Equatorial Guinea | GNQ | Central | Non-Gavi | Non-Gavi | No | No | No | 0 |
| Eritrea | ERI | E. & Southern | Gavi | Gavi, non-conflict-affected | No | Yes | Yes | **1** |
| Eswatini | SWZ | E. & Southern | Non-Gavi | Non-Gavi | No | No | No | **2** |
| Ethiopia | ETH | E. & Southern | Gavi | Gavi, conflict-affected | Yes | No | Yes | **3** |
| Gabon | GAB | Central | Non-Gavi | Non-Gavi | No | No | No | 0 |
| The Gambia | GMB | West | Gavi | Gavi, non-conflict-affected | No | No | No | 0 |
| Ghana | GHA | West | Gavi | Gavi, non-conflict-affected | No | No | No | **1** |
| Guinea | GIN | West | Gavi | Gavi, non-conflict-affected | No | No | No | **1** |
| Guinea-Bissau | GNB | West | Gavi | Gavi, non-conflict-affected | No | Yes | Yes | **1** |
| Kenya | KEN | E. & Southern | Gavi | Gavi, non-conflict-affected | No | No | No | **1** |
| Lesotho | LSO | E. & Southern | Gavi | Gavi, non-conflict-affected | No | No | No | 0 |
| Liberia | LBR | West | Gavi | Gavi, non-conflict-affected | No | No | No | **1** |
| Madagascar | MDG | E. & Southern | Gavi | Gavi, non-conflict-affected | No | No | No | **1** |
| Malawi | MWI | E. & Southern | Gavi | Gavi, non-conflict-affected | No | No | No | **2** |
| Mali | MLI | West | Gavi | Gavi, conflict-affected | Yes | No | Yes | **3** |
| Mauritania | MRT | West | Gavi | Gavi, non-conflict-affected | No | No | No | **1** |
| Mauritius | MUS | E. & Southern | Non-Gavi | Non-Gavi | No | No | No | 0 |
| Mozambique | MOZ | E. & Southern | Gavi | Gavi, conflict-affected | Yes | No | Yes | **1** |
| Namibia | NAM | E. & Southern | Non-Gavi | Non-Gavi | No | No | No | **1** |
| Niger | NER | West | Gavi | Gavi, conflict-affected | Yes | No | Yes | **3** |
| Nigeria | NGA | West | Gavi | Gavi, conflict-affected | Yes | No | Yes | **4** |
| Rwanda | RWA | E. & Southern | Gavi | Gavi, non-conflict-affected | No | No | No | **1** |
| São Tomé and Príncipe | STP | Central | Gavi | Gavi, non-conflict-affected | No | Yes | Yes | 0 |

| Country | ISO3 | Subregion | Gavi | Financing–conflict group | Confl. | Frag. | Br.-FCS | Events |
|---|---|---|---|---|---|---|---|---|
| Senegal | SEN | West | Gavi | Gavi, non-conflict-affected | No | No | No | **1** |
| Seychelles | SYC | E. & Southern | Non-Gavi | Non-Gavi | No | No | No | **2** |
| Sierra Leone | SLE | West | Gavi | Gavi, non-conflict-affected | No | No | No | **1** |
| South Africa | ZAF | E. & Southern | Non-Gavi | Non-Gavi | No | No | No | **2** |
| South Sudan | SSD | E. & Southern | Gavi | Gavi, conflict-affected | Yes | No | Yes | **3** |
| Tanzania | TZA | E. & Southern | Gavi | Gavi, non-conflict-affected | No | No | No | 0 |
| Togo | TGO | West | Gavi | Gavi, non-conflict-affected | No | No | No | **2** |
| Uganda | UGA | E. & Southern | Gavi | Gavi, non-conflict-affected | No | No | No | **1** |
| Zambia | ZMB | E. & Southern | Gavi | Gavi, non-conflict-affected | No | No | No | **3** |
| Zimbabwe | ZWE | E. & Southern | Gavi | Gavi, non-conflict-affected | No | Yes | Yes | 0 |

**Angola: non-Gavi in the primary analysis (its recorded events occurred in 2025); examined as Gavi-eligible in sensitivity analysis (Supplementary Table S5). Confl. = World Bank FY26 conflict-affected category; Frag. = institutional and social fragility category; Br.-FCS = broader fragile and conflict-affected situation (either category). Reconciliation (n = 47): non-Gavi 11; Gavi-eligible non-conflict-affected 26; Gavi-eligible conflict-affected 10. World Bank FY26 conflict-affected = 10; institutional/social fragility = 8; Gavi-eligible broader-FCS group = 18 (36 events). Because conflict-affected status was nested within Gavi eligibility, the three-level grouping avoids interpreting the conflict-affected contrast as a simple Gavi versus non-Gavi comparison.*

### Supplementary Table S4. Baseline introduction-opportunity index

Baseline introduction opportunity was defined as the count (0–7) of seven recommended products or schedule components not yet introduced nationally by the end of 2022, using WHO/UNICEF national introduction-year data. The index adjusts for differential room to introduce, so that the financing–conflict gradient is not an artefact of how many recommended vaccines a country had still to adopt.

| Component counted in the index (not yet introduced by end-2022) | Type |
|---|---|
| Pneumococcal conjugate vaccine (PCV) | Antigen |
| Rotavirus vaccine | Antigen |
| Hepatitis B birth dose | Schedule component |
| Second dose of inactivated poliovirus vaccine (IPV2) | Schedule component |
| Second-dose measles-containing vaccine (MCV2) | Schedule component |
| Rubella-containing vaccine | Antigen |
| Human papillomavirus (HPV) vaccine | Antigen |

*Targeted products (malaria, typhoid and meningococcal vaccines) were excluded from the index by design. The per-country index for all 47 Member States is provided in the deposited Supplementary Workbook (worksheet S4). In the baseline-opportunity-adjusted model the Gavi-eligible conflict-affected association persisted (IRR 2.00, 95% CI 1.36–2.94), and remaining introduction opportunity was not itself associated with event counts (IRR 0.99, 95% CI 0.90–1.08; p = 0.78), indicating the gradient was not explained by differential room to introduce.*

### Supplementary Table S5. Primary and sensitivity model outputs

All models are country-level ecological count models (N = 47) with HC1 robust standard errors. Reported p values are descriptive compatibility measures rather than confirmatory tests.

| Analysis | Countries / events | Reference group | Non-Gavi IRR (95% CI; p) | Conflict-affected / broader-FCS IRR (95% CI; p) |
|---|---|---|---|---|
| Primary three-level Poisson | 47 / 72 | Gavi non-conflict-affected | 1.00 (0.54–1.87; 0.99) | 1.97 (1.38–2.81; <0.001) |
| New-antigen events only | 47 / 48 | Gavi non-conflict-affected | — | 2.36 (1.43–3.91; 0.001) |
| Non-new-antigen events only | 47 / 24 | Gavi non-conflict-affected | — | 1.13 (0.37–3.41; 0.84) |
| Malaria-vaccine events excluded | 47 / 51 | Gavi non-conflict-affected | — | 1.77 (1.06–2.97; 0.030) |
| January–June 2026 excluded | 47 / 65 | Gavi non-conflict-affected | — | 2.24 (1.51–3.33; <0.001) |
| Angola reclassified Gavi-eligible | 47 / 72 | Gavi non-conflict-affected | — | 1.93 (1.36–2.73; <0.001) |
| Baseline-opportunity adjusted | 47 / 72 | Gavi non-conflict-affected | — | 2.00 (1.36–2.94; <0.001) |
| Remaining-opportunity covariate† | 47 / 72 | — | — | 0.99 (0.90–1.08; 0.78) |
| Negative-binomial (overdispersion check) | 47 / 72 | Gavi non-conflict-affected | — | 1.97 (concordant; full output in code) |
| Leave-one-country-out range | 46 / varies | Gavi non-conflict-affected | — | 1.84–2.10 |
| Broader World Bank FCS definition | 47 / 72 | Gavi non-broader-FCS | 1.04 (0.54–1.99; 0.903) | 1.64 (1.08–2.47; 0.019) |

*Primary group means: non-Gavi 14/11 = 1.27; Gavi-eligible non-conflict-affected 33/26 = 1.27; Gavi-eligible conflict-affected 25/10 = 2.50. Broader-FCS group means: non-Gavi 14/11 = 1.27; Gavi-eligible non-broader-FCS 22/18 = 1.22; Gavi-eligible broader-FCS 36/18 = 2.00. Primary-model diagnostics: Pearson $\chi^2$/df 0.75; deviance/df 0.91 (no overdispersion). †For this row the estimate refers to the remaining-opportunity covariate, not the conflict-affected contrast. IRR = incidence rate ratio; FCS = fragile and conflict-affected situations.*

### Supplementary Table S6. EPI manager survey indicators, domains and denominators

Two EPI manager survey waves were analysed separately as programme-reported implementation data and were not pooled. Country-collapsing used a predefined respondent hierarchy (National EPI Manager > Deputy > technical officer > other).

| Indicator (response option counted) | Wave | Domain | n / N | % |
|---|---|---|---|---|
| Ad hoc prioritisation process | 2023 | Decision governance | 20 / 37 | 54.1 |
| Limited resources or data as main challenge | 2023 | Decision governance | 19 / 37 | 51.4 |
| Willing to adopt a structured MCDA tool | 2023 | Decision governance | 30 / 37 | 81.1 |
| Financing and sustainability as top challenge | 2025 | Financing | 28 / 35 | 80.0 |
| Donor-related delay to introduction plans | 2025 | Financing | 7 / 35 | 20.0 |
| Product or formulation switch, past 5 years | 2025 | Product management | 20 / 35 | 57.1 |
| Preparatory activities not started (of 2026 planners) | 2025 | Preparation | 9 / 25 | 36.0 |
| Recent post-introduction evaluation | 2025 | Accountability | 13 / 35 | 37.1 |

*The 2026-preparation denominator is the 25 countries planning a 2026 introduction. Findings are descriptive programme intelligence and should not be read as population prevalence or as a longitudinal trend. Full instruments are available from the corresponding author subject to institutional clearance. MCDA = multi-criteria decision analysis.*

### Supplementary Table S7. Immunization League Table (ILT): composition, interpretation and concurrent-indicator correlations

| Attribute | Description | Interpretation / limitation |
|---|---|---|
| Nature | Internal WHO African Region benchmarking and prioritisation tool to identify immunity and equity gaps and target differentiated support | Not an externally validated readiness index |
| Use here | Concurrent programme-performance marker (score and band) | Concurrent and descriptive; does not measure readiness or predict outcomes |
| Composition | Composite of immunisation-performance elements including DTP3 coverage and zero-dose burden | Partial non-independence with DTP3 and zero-dose analyses |
| Banding | Very-low / low / moderate / high performance bands | Relative regional rankings, not absolute thresholds |
| Source / period | WHO African Region programme dashboard, 2025 | Internal document; not publicly available |

#### Concurrent system-performance associations (descriptive)

Countries in low or very-low ILT bands were more likely to record two or more events than countries in moderate or high bands (15 of 24, 62.5%, versus 7 of 23, 30.4%; Fisher's exact $p = 0.041$). Event counts were inversely associated with the UHC service coverage index. Because the ILT incorporates DTP3 and zero-dose information, its association with event counts is partly mechanical. These exploratory correlations did not survive multiplicity adjustment and describe ecological co-occurrence, not temporal prediction, implementation readiness or causal effect.

### Supplementary Figure S1. Event composition by period and vaccine or platform

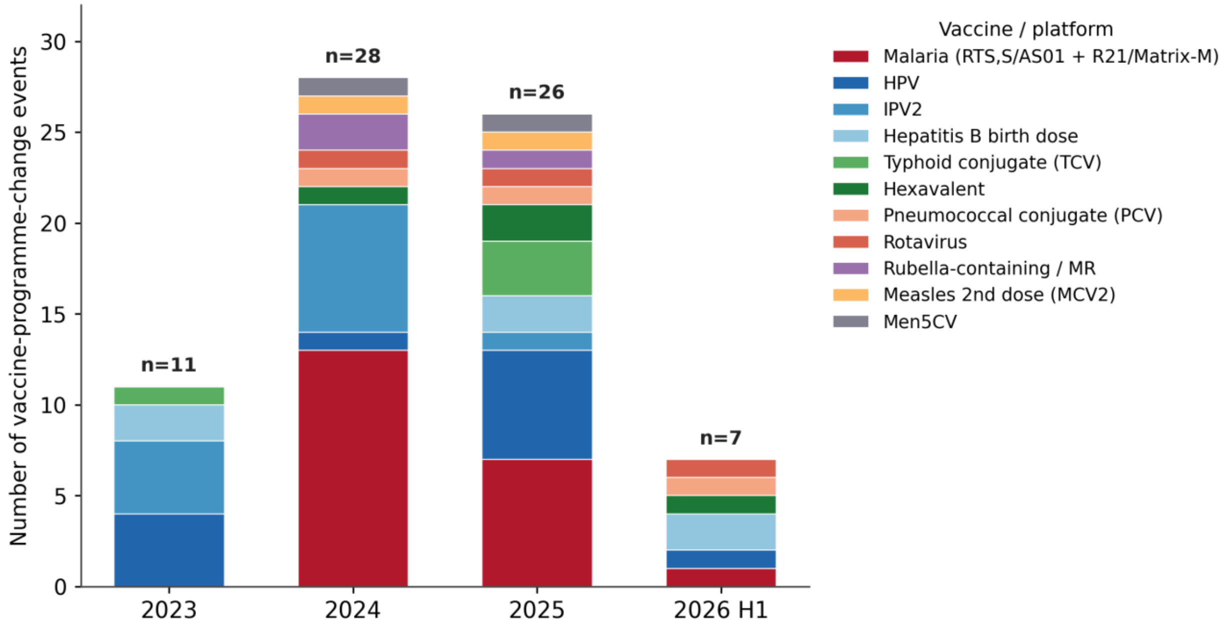


*Stacked counts of the 72 vaccine-programme-change events by calendar period and vaccine or platform. Period totals are shown above each bar. The 2026 H1 column covers January–June 2026 only and is not annualised. Event activity clustered in 2024–2025 (54 of 72 events, 75.0%), with malaria vaccines the largest single contributor (21 events).*

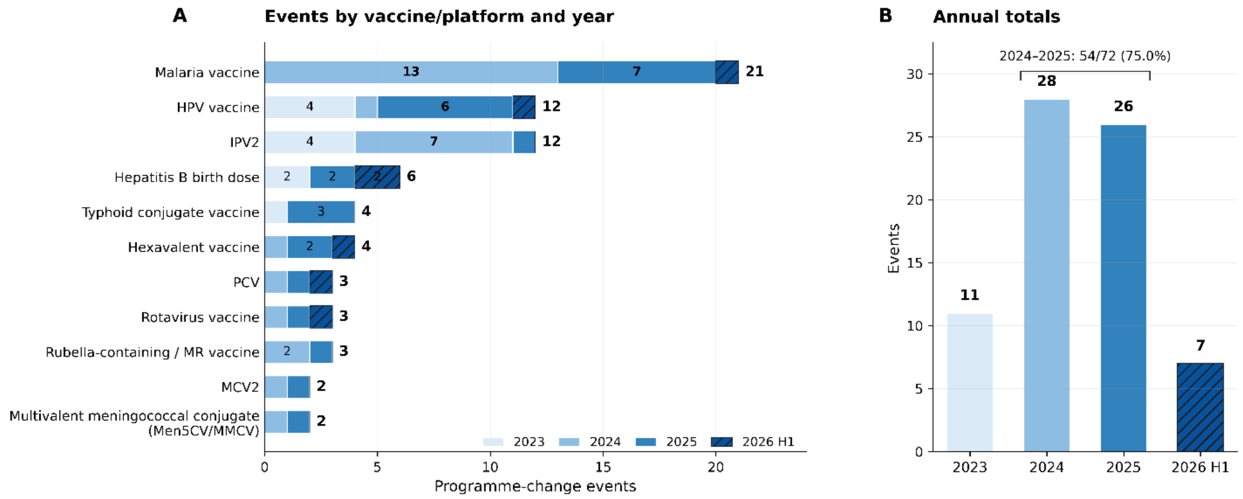


### Supplementary Figure S2. Robustness and leave-one-country-out influence analysis

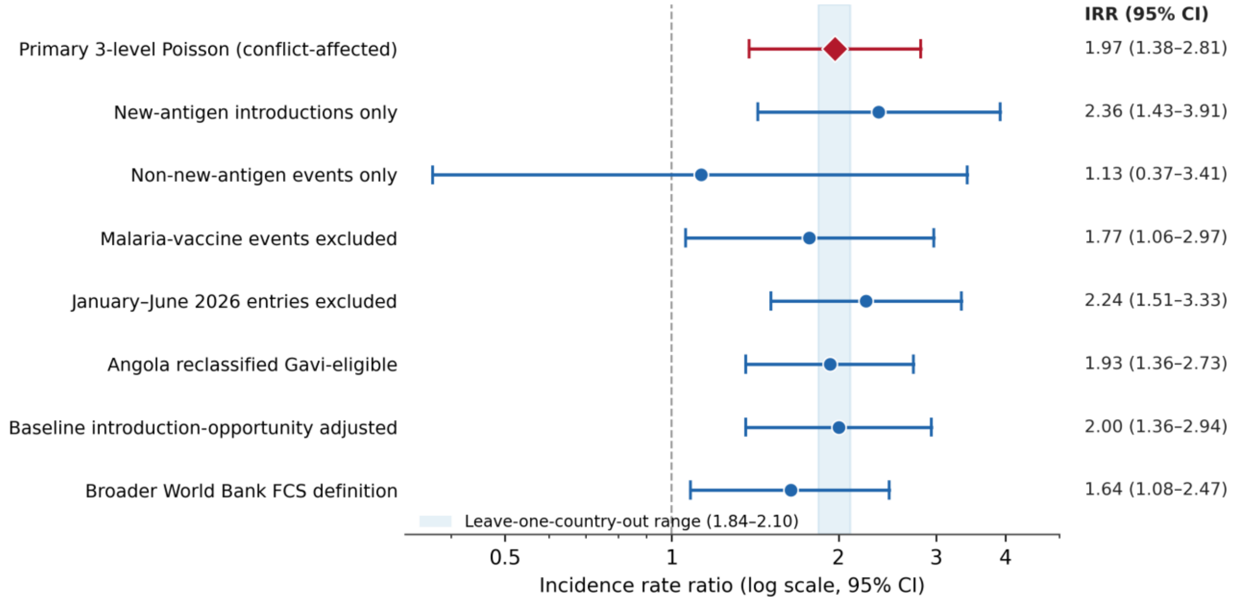


*Incidence rate ratios (IRRs) for the Gavi-eligible conflict-affected contrast across model specifications, on a logarithmic scale with 95% confidence intervals. The reference group is Gavi-eligible non-conflict-affected countries for all rows except the final row, which uses Gavi-eligible non-broader-FCS countries. The red diamond marks the primary estimate (IRR 1.97, 95% CI 1.38–2.81); the dashed line marks no association (IRR 1.0). The shaded vertical band shows the leave-one-country-out range (1.84–2.10), indicating that no single country drove the primary estimate. The association attenuated but persisted after excluding malaria-vaccine events (IRR 1.77) and under the broader World Bank FCS definition (IRR 1.64).*

### Supplementary File S1. STROBE and RECORD reporting

Reporting followed the STROBE statement for observational studies and the RECORD extension for studies using routinely collected health data. A completed item-by-item STROBE/RECORD checklist accompanies the submission as a separate file.

Key mappings: study design and setting (Methods 2.1); eligibility and event definition (Methods 2.2; Table S1); data sources, linkage and cleaning (Methods 2.3; RECORD items 6.1–6.3); variables and classification (Methods 2.4; Table S3); statistical methods, robustness and bias handling (Methods 2.6; Table S5; Figure S2); descriptive and model results (Results 3.1–3.7; main Tables 1–3; main Figures 1–3); and limitations of routinely collected data (Discussion, Strengths and limitations). RECORD items on database population, linkage and code lists are addressed by the event-inclusion rules (Table S1) and the country master file (Table S3; Supplementary Workbook).

### Supplementary File S2. Reproducibility and analysis code

Analyses used Python 3.12 (numpy 1.26, pandas 2.2, statsmodels 0.14) and R 4.4. The primary model was a Poisson regression of the country-level event count on the three-level financing–conflict group with HC1 robust standard errors and the Gavi-eligible non-conflict-affected group as the reference; a negative-binomial model was fitted as an overdispersion check. Group-mean confidence intervals used a nonparametric bootstrap with 10,000 resamples, resampling countries within group, with random seed 2026.

The repository contains the analysis scripts that generate main Tables 1–3 and Figures 1–3, the supplementary tables and figures, the data dictionary and the country master file, together with a reproducibility README. The repository will be deposited publicly on acceptance, subject to institutional clearance.
