## Supplementary material for "Vaccine introductions in the WHO African Region, 2023–26: a country-level ecological analysis by Gavi eligibility and conflict-affected status": STROBE RECORD checklist

### STROBE and RECORD reporting checklist

This checklist maps the manuscript against the STROBE statement for observational studies and the RECORD extension for routinely collected health data. Item numbers follow STROBE (2007) with relevant RECORD (2015) extensions.

| Section / topic | Item | Reported | Location |
|---|---|---|---|
| **Title and abstract** | STROBE 1; RECORD 1.1-1.3 | Study design, setting, population, data sources and main results are stated in the title and structured abstract. | Title; Abstract |
| **Introduction - background/rationale** | STROBE 2 | Scientific background, rationale and the limitation of launch-status monitoring are described. | Introduction |
| **Objectives** | STROBE 3 | Primary and secondary objectives are stated, including timing, portfolio composition, financing-conflict grouping, concurrent system indicators and EPI manager-reported constraints. | Introduction, final paragraph |
| **Study design** | STROBE 4 | Descriptive country-level ecological design is stated. | Methods 2.1 |
| **Setting** | STROBE 5 | Setting, locations and observation period are reported: all 47 WHO African Region Member States, January 2023 to June 2026. | Methods 2.1 |
| **Participants / units of analysis** | STROBE 6; RECORD 6.1-6.3 | The country is the unit of analysis; all 47 Member States are included; linkage, validation and classification procedures are described. | Methods 2.1-2.4; Supplementary Tables S1-S3 |
| **Variables** | STROBE 7 | Outcome, exposure grouping and concurrent indicators are defined: event count, financing-conflict group, system-performance indicators and EPI manager survey indicators. | Methods 2.2-2.5; Supplementary Tables S1-S3 and S6-S7 |
| **Data sources / measurement** | STROBE 8; RECORD 7.1 | Data sources, event eligibility, classification, country master variables, survey measurement and indicator limitations are reported. | Methods 2.2-2.5; Supplementary Tables S1-S3 and S6-S7; Data availability statement |
| **Bias** | STROBE 9 | Ecological fallacy, concurrency, sparse models, multiplicity, non-response, event heterogeneity, indicator non-independence and author positionality are addressed. | Methods 2.6; Discussion, Strengths and limitations |
| **Study size** | STROBE 10 | Full regional census of 47 Member States is reported; no sampling calculation was required. | Methods 2.1 |
| **Quantitative variables** | STROBE 11 | Grouping variables, event counts, group means, baseline introduction-opportunity proxy and concurrent indicators are described. | Methods 2.4 and 2.6; Table 2; Supplementary Tables S3-S4 and S7 |
| **Statistical methods** | STROBE 12 | Poisson regression with HC1 robust standard errors, bootstrap group-mean confidence intervals, overdispersion checks, sensitivity analyses, exploratory correlations and no multiplicity correction are described. | Methods 2.6; Table 2; Supplementary Tables S4-S5; Supplementary Figure S2; Supplementary File S2 |
| **Data access, linkage and cleaning** | RECORD 12.1-12.3 | Event reconciliation, standardisation, classification, de-duplication, validation and reproducibility information are described. | Methods 2.2-2.3; Supplementary Tables S1-S3; Supplementary File S2; Data availability statement |
| **Descriptive data** | STROBE 13-14; RECORD 13.1 | Counts by year, event type, vaccine/platform, financing-conflict group, subregion and EPI manager survey denominators are reported. | Results 3.1-3.6; Tables 1-3; Supplementary Tables S2-S3 and S6-S7 |
| **Outcome data / main results** | STROBE 15-16 | Event counts, group means, incidence rate ratios and 95% confidence intervals are reported, with group-mean CIs distinguished from model-based robust IRR CIs. | Results 3.3-3.7; Figure 2; Table 2; Supplementary Tables S2 and S5 |
| **Other analyses** | STROBE 17 | New-antigen, non-new-antigen, malaria-exclusion, partial-year exclusion, Angola reclassification, baseline-opportunity, broader-FCS and leave-one-country-out analyses are reported. | Results 3.4 and 3.7; Supplementary Table S5; Supplementary Figure S2 |
| **Key results** | STROBE 18 | Main findings are summarised with reference to the study objectives and bounded by the ecological design. | Discussion, opening paragraphs |
| **Limitations** | STROBE 19; RECORD 19.1 | Limitations include unweighted event counts, ecological design, sparse models, concurrency, non-independent indicators, non-response, self-reporting, event heterogeneity, Men5CV classification and author positionality. | Discussion, Strengths and limitations |
| **Interpretation** | STROBE 20 | Interpretation is cautious, non-causal and framed as an implementation-assurance and distributional-risk signal rather than causal evidence. | Discussion |
| **Generalisability** | STROBE 21 | Scope of inference is restricted to the WHO African Region and distinguishes World Bank conflict-affected status from the broader FCS sensitivity definition. | Methods 2.4; Discussion, Strengths and limitations |
| **Funding** | STROBE 22 | Funding statement is provided. | Declarations, Funding |
| **Data availability / accessibility** | RECORD 22.1 | Data availability, public-source indicators, internal programme-document restrictions, planned repository deposition and reproducibility-file information are described. | Declarations, Data availability; Supplementary File S2 |

**Conclusion.** All STROBE items and relevant RECORD extensions for routinely collected health data are addressed in the manuscript and supplementary files. This checklist intentionally follows the manuscript primary classification as financing-conflict; broader FCS is mapped only as a sensitivity analysis.
